# Postnatal CMV acquisition risk is not increased among children who are HIV-exposed, uninfected versus HIV-unexposed in the setting of optimized maternal ART

**DOI:** 10.64898/2026.10.27.26365941

**Authors:** Emily R. Begnel, Mi-Suk Kang Dufour, Kerusha Govender, Jing Hu, Elisabeth McClymont, Ednah Ojee, Prestone Owiti, Judith Adhiambo, Eliza Mabele, Isabelle Boucoiran, Fatima Kakkar, Bhavna H. Chohan, Dara A. Lehman, John Kinuthia, Dalton Wamalwa, Soren Gantt, Jennifer Slyker

## Abstract

Historically, children who are HIV-exposed, uninfected (CHEU) had higher incidence and earlier postnatal acquisition of cytomegalovirus (CMV) than children who are HIV-unexposed. We assessed whether this difference persists in the current era of universal antiretroviral therapy (ART). Women without HIV and women living with HIV (WLHIV) on ART for ≥6 months were recruited in pregnancy in Nairobi, Kenya and followed weekly with their infant for 12 months postpartum; 89% of WLHIV initiated ART pre-conception. Log_10_-CMV DNA levels were measured in infant urine, oral swabs, and plasma, and in maternal breast milk, oral swabs, and vaginal swabs. Infant plasma collected at age ≥12 months was tested for CMV IgG antibodies. Kaplan-Meier survival analysis and Cox proportional hazards regression were used to compare CMV acquisition by HIV exposure and assess correlates of CMV infection. Of 65/72 (90%) infants with ascertainable CMV status and timing of acquisition (42% CHEU), 47 (72%) acquired CMV. WLHIV had higher mean CMV DNA levels than women without HIV in breast milk (4.17-log vs 3.12-log, p<0.001) and oral swabs (1.10-log vs 0.78-log, p<0.001). Though higher breast milk and oral swab CMV DNA levels were associated with increased hazard of CMV among all infants (adjusted hazard ratio [aHR]=2.80, 95% CI: 1.77-4.43 and aHR=1.35, 95% CI: 1.14-1.60), HIV exposure was not significantly associated with CMV acquisition (aHR=1.31, 95% CI: 0.71-2.42). Hazard of CMV was lower among CHEU that received cotrimoxazole prophylaxis (aHR=0.19, 95% CI: 0.074-0.47) and whose mothers received a recommended first line ART regimen postpartum (aHR=0.37, 95% CI: 0.15-0.93). Together, these findings suggest that risk of CMV infection among CHEU is modulated by level of maternal mucosal CMV shedding, maternal ART regimen, and cotrimoxazole in this setting of optimized, pre-conception ART. Breast milk remains an important source of postnatal CMV acquisition for all infants.

## INTRODUCTION

Cytomegalovirus (CMV) infection is common worldwide, with seroprevalence as high as 100% in low- and middle-income countries [1,2]. Primary CMV infection establishes latency, resulting in lifelong infection [3] that periodically reactivates, particularly with inflammation and immune deficiency [3,4]. Reinfection with new strains occurs frequently despite a robust adaptive immune response to initial infection [5]. Following infection or reactivation, infectious CMV is shed at mucosal surfaces and is transmitted through direct contact with body fluids [6,7].

Globally, 40-60% of infants with a CMV-seropositive mother will acquire CMV by 12 months, though this proportion is often greater in populations with high seroprevalence [8,9]. Breastfeeding is a major route of CMV transmission. CMV reactivation in breast milk occurs in virtually all CMV-seropositive lactating women [10,11], resulting in transmission via infected breast milk cells and cell-free virus [7,12]. Though CMV is the most common congenital infection, occurring in roughly 1% of all live births, and represents a leading cause of hearing loss and neurodevelopmental disability in children [5,13], most postnatal CMV infections in healthy children are asymptomatic [3,4]. However, CMV is highly immunomodulatory and indirect effects of CMV infection, including an excess of non-infectious morbidity and mortality, are increasingly recognized [14,15]. In pediatric populations, CMV infection has been shown to alter immune responses to other pathogens and vaccines, and exacerbate conditions contributing to morbidity and mortality such as malnutrition and pneumonia [16–20]. While preventing CMV transmission is an important public health priority, no effective CMV vaccine is yet available [21].

Historically, children who are HIV-exposed, uninfected (CHEU) have had higher incidence of congenital CMV and earlier acquisition of postnatal CMV infection than children who are HIV-unexposed (CHU), and these differences persisted through the early antiretroviral therapy (ART) era [9,22]. Contributing factors include immunologic differences in CHEU versus CHU [23,24] and increased exposure to CMV shedding in breast milk and saliva from women living with HIV (WLHIV) [9,25]. ART-mediated immune recovery among mothers may reduce or delay postnatal CMV transmission to CHEU. However, there are few data comparing the incidence of postnatal CMV acquisition between CHEU and CHU in the current setting of universal maternal ART. To address this gap, we assessed whether the timing and incidence of CMV acquisition differed between CHEU born to WLHIV who initiated modern ART pre-conception or in early pregnancy and their CHU peers. Additionally, we defined correlates of CMV acquisition among CHEU and CHU and compared breast milk CMV DNA levels by maternal HIV status.

## METHODS

### Study Population

This analysis was nested in the Linda Kizazi Study, a prospective cohort of mother-infant pairs in Nairobi, Kenya followed from delivery to 24 months postpartum as previously described [26]. WLHIV and women without HIV were eligible if they were 28-42 weeks pregnant, 18-40 years old, planned to breastfeed, did not have a planned Caesarean section, were not diagnosed with a serious medical condition, and had received ≥6 months of ART if living with HIV. All participants received a home visit 4 days postpartum, then visits at the study clinic approximately every 3 months between week 6 and month 24. At enrollment, participants were offered participation in additional weekly home visits between delivery and 12 months; mother-infant pairs who self-selected to receive these visits were included in this nested analysis. If there were twins, only the firstborn infant was included in analyses. Participants provided written informed consent for themselves and their infant. All Linda Kizazi Study procedures and data analyses were approved by the Kenyatta National Hospital-University of Nairobi Ethics and Research Committee (P472/07/2018) and the University of Washington Institutional Review Board (STUDY00004006).

### Data & Specimen Collection

Participants’ sociodemographic characteristics and health history were recorded at enrollment. At all visits, study clinicians collected data on maternal and infant health, medications, and infant feeding. At the weekly visits, clinicians collected oral swabs from mothers and infants, and samples of infant urine and maternal breast milk. Saliva was collected with Dacron swabs ≥1 hour after breastfeeding or ≥15 minutes after eating using a standardized procedure, then stored at room temperature in 1ml of premixed buffer prepared by the study laboratory (solution of 18.65g KCl, 500g ACS Reagent, 25ml Tris HCl 1M [pH 8.0], 125ml EDTA 0.5M, 25ml IGEPAL CA_630, and 325ml sterile water, diluted in a 1:4 ratio with additional sterile water) [9]. One milliliter of solution was aliquoted into a 2ml cryovial that was sealed and stored at room temperature until use for sample collection. Infant urine was collected into an adhesive collection bag, aliquoted into 2ml vials, and stored at −80°C without preservatives. Mothers self-collected 10ml of breast milk from each breast; to obtain a “clean-catch” sample, they first cleaned each breast with an alcohol wipe, then expressed and discarded the first drops of breast milk. Breast milk samples were centrifuged at 931 x g at 4°C for 25 minutes to separate and remove the lipid layer, aliquoted into 2ml vials, and stored at −80°C without preservatives. Additionally, a maternal vaginal sample was collected at enrollment with a flocked swab stored in 3ml of universal viral transport media (Universal Viral Transport System, Becton, Dickinson and Company, USA) at −80°C. At clinic visits, clinicians collected oral swabs from mothers and infants, and 4ml of whole blood from infants into EDTA vacutainers. Plasma samples were prepared from whole blood centrifuged at 754 x g at room temperature for 10 minutes, aliquoted into 2ml vials, and stored at −80°C.

### CMV Testing

Quantitative polymerase chain reaction (qPCR) was used to measure CMV DNA levels, which were log_10_-transformed for analysis. For urine, oral swabs, breast milk, and vaginal swabs, CMV DNA was extracted from 500µl of sample using the AltoStar® Purification Kit 1.5 and qPCR was performed using the AltoStar® CMV PCR Kit 1.5 via the AltoStar® Automation System AM16 according to manufacturer instructions (Altona Diagnostics, Germany). The limit of detection was 2.85-log International Units (IU)/ml (95% CI: 2.62-3.21) for urine, 2.60-log IU/ml (95% CI: 2.43-2.93) for oral swabs, 2.28-log IU/ml (95% CI: 2.07-2.63) for breast milk, and 2.33-log IU/ml (95% CI: 2.14-2.64) for vaginal swabs; results with a quantifiable viral load were considered positive [27]. For plasma, CMV DNA was extracted from 20-200µl of sample and qPCR was performed as previously described [28]; the limit of detection was 1.70-log IU/ml. CMV IgG antibodies were measured in infant plasma collected between ages 9-15 months, to exclude maternal transfer of antibodies, using the IBL CMV IgG ELISA kit according to manufacturer instructions (Immuno-Biological Laboratories Inc., USA); an antibody index of >0.5 was considered positive.

No infants acquired HIV during study follow-up; all infants born to WLHIV were classified as CHEU. Infants were classified as CMV infected if they had a positive serology at ≥12 months or, if serology was not available, by positive blood PCR and/or sustained shedding in either urine or oral swabs. Sustained shedding was defined as at least 3 consecutive PCR positive results in a sample type, above a threshold of ≥3-log in oral swabs or ≥2-log in urine. Estimated time of infection was defined as the date of either the first indication of sustained shedding or positive serology/plasma PCR result.

### Statistical Analyses

Kaplan-Meier survival analysis was used to assess the probability and timing of CMV acquisition. Time at risk was defined as months between birth and the time of infection or, if CMV-negative, the collection date of the last sample tested for CMV DNA. Incidence was calculated as the number of infections per 100 person-months at risk. Cox proportional hazards regression with robust standard errors was used to measure the association between potential risk factors previously shown or hypothesized to be associated with CMV infection, including: HIV exposure; infant sex; feeding mode; living with siblings <5 years old; household crowding (>3 people/room in house); sharing a toilet with other households; detectable CMV DNA in the maternal vaginal swab at enrollment; and log_10_-CMV DNA levels in maternal breast milk and oral swabs at each visit. Among CHEU, we also evaluated maternal enrollment CD4 count and ART regimen, visit-level maternal postpartum ART regimen, and infant visit-level cotrimoxazole prophylaxis. For maternal ART, we compared exposure to the contemporary recommended first-line ART regimen (TDF+3TC+EFV or TDF+3TC+DTG) versus exposure to alternative first-line or secondary regimens.

Feeding mode was defined as exclusive breastfeeding (infant received only breast milk and prescribed medication), mixed feeding (breast milk and other foods, liquids, and/or medications), and weaned (only foods and liquids other than breast milk). All participants included in this analysis initiated breastfeeding at delivery and breastfed continually (exclusively or as mixed feeding) until either their infant was weaned or their follow-up ended; accordingly, all visits without breastfeeding exposure occurred after the infant was weaned.

Models were adjusted, as applicable, for HIV exposure, infant sex, and household crowding, as these variables were determined *a priori* to be potential confounders. Proportions were compared using Chi-square tests, medians were compared using Wilcoxon rank-sum tests, and means were compared using t-tests. Descriptive plots of CMV levels were created in R (version 4.3.3). All analyses were conducted in Stata (version 17) with a significance level of 0.05.

## RESULTS

### Participant Characteristics

Of 211 women enrolled in the Linda Kizazi Study, 72 (34%) elected to receive weekly home visits for 12 months postpartum, of which 28 (39%) were living with HIV. Follow-up was similar between CHEU and CHU (median 42 vs 40 visits, p=0.318). CMV DNA levels were measured in: 1873 infant urine, 2915 infant oral swab, 2223 maternal breast milk, and 2686 maternal oral swab samples collected approximately weekly for up to one year postpartum; 72 maternal vaginal swab samples collected at enrollment; and 180 infant plasma samples collected at quarterly clinic visits.

Compared to women without HIV, fewer WLHIV had other children <5 years old (61% vs 32%, p=0.016; Table 1). Though a larger proportion of WLHIV than women without HIV experienced crowding (75% vs 61%), this difference was not statistically significant (p=0.231). Sharing a toilet with other households was common (90%). Most infants were delivered vaginally (96%), and adverse birth outcomes were rare—one CHEU was born preterm (<37 weeks gestation), and one CHEU and one CHU were born with low birthweight (<2.5 kg). A greater proportion of CHU than CHEU were male (64% vs 39%, p=0.043). More CHEU than CHU were exclusively breastfed (EBF) for 6 months (82% vs 55%, p=0.028) as recommended by the WHO [29], though duration of any breastfeeding was longer among CHU (median 18 vs 12 months, p<0.001).

**Table 1.** Selected participant characteristics.

|  |  | All |  | HIV-unexposed<br>uninfected |  | HIV-exposed<br>uninfected |
| --- | --- | --- | --- | --- | --- | --- |
|  | N | Median (IQR) or n (%) | N | Median (IQR) or n (%) | N | Median (IQR) or n (%) |
| **Maternal Characteristics** |  |  |  |  |  |  |
| Age (years) at enrollment | 72 | 26 (24-30) | 44 | 25 (23-30) | 28 | 27 (24-31) |
| Married or steady partner | 72 | 68 (94.4) | 44 | 43 (97.7) | 28 | 25 (89.3) |
| Highest education level | 72 |  | 44 |  | 28 |  |
| Secondary or higher |  | 42 (58.3) |  | 27 (61.4) |  | 15 (53.6) |
| Primary or lower |  | 30 (41.7) |  | 17 (38.6) |  | 13 (46.4) |
| Employed | 72 | 24 (33.3) | 44 | 14 (31.8) | 28 | 10 (35.7) |
| Weekly income (USD)<sup>1</sup> | 16 | 15.00 (8.75-27.00) | 11 | 15.00 (5.00-30.00) | 5 | 12.50 (10.00-20.00) |
| Lifetime number of pregnancies<sup>2</sup> | 72 | 2 (2-3) | 44 | 2 (2-3) | 28 | 3 (2-4) |
| Number of living children<sup>3</sup> | 60 | 2 (1-2) | 35 | 1 (1-2) | 25 | 2 (1-3) |
| **Sociodemographics** |  |  |  |  |  |  |
| Home roofing material | 72 |  | 44 |  | 28 |  |
| Metal |  | 54 (75.0) |  | 32 (72.7) |  | 22 (78.6) |
| Concrete |  | 17 (23.6) |  | 12 (27.3) |  | 5 (17.9) |
| Other |  | 1 (1.4) |  | 0 (0.0) |  | 1 (3.6) |
| Experienced crowding<br>(>3 people/room) in home | 72 | 48 (66.7) | 44 | 27 (61.4) | 28 | 21 (75.0) |
| Any siblings <5 years<sup>3</sup> | 72 | 36 (50.0) | 44 | 27 (61.4) | 28 | 9 (32.1) |
| Number of siblings <5 years<sup>4</sup> | 36 | 1 (1-1) | 27 | 1 (1-1) | 9 | 1 (1-1) |
| Toilet is shared with other<br>households | 72 | 65 (90.3) | 44 | 39 (88.6) | 28 | 26 (92.9) |
| Home has running water | 72 | 19 (26.4) | 44 | 10 (22.7) | 28 | 9 (32.1) |
| Home has electricity | 72 | 70 (97.2) | 44 | 43 (97.7) | 28 | 27 (96.4) |
| **Infant Characteristics** |  |  |  |  |  |  |
| Male sex assigned at birth | 72 | 39 (54.2) | 44 | 28 (63.6) | 28 | 11 (39.3) |
| Vaginal birth | 69 | 66 (95.7) | 43 | 41 (95.4) | 26 | 25 (96.2) |
| Gestational age at birth (weeks) | 68 | 38 (38-40) | 43 | 38 (38-40) | 25 | 38 (38-39) |
| Birthweight (kg) | 68 | 3.3 (3.0-3.6) | 42 | 3.4 (3.2-3.7) | 26 | 3.2 (3.0-3.5) |
| Ever breastfed | 72 | 72 (100.0) | 44 | 44 (100.0) | 28 | 28 (100.0) |
| Exclusively breastfed for 6 months<sup>5</sup> | 65 | 43 (66.2) | 38 | 21 (55.3) | 27 | 22 (81.5) |
| Months of exclusive breastfeeding<sup>5</sup> | 65 | 6 (5-6) | 38 | 6 (5-6) | 27 | 6 (6-6) |
| Months of any breastfeeding<sup>6</sup> | 54 | 15 (12-19) | 33 | 18 (15-20) | 21 | 12 (11-13) |

IQR = Interquartile range. USD = US dollars. <sup>1</sup> If employed. <sup>2</sup> Includes enrolled infant. <sup>3</sup> If ≥1 pregnancy prior to pregnancy at enrollment. <sup>4</sup> If mother reporting having other children <5 years at time of enrollment. <sup>5</sup> Among mother-infant pairs followed for ≥6 months. <sup>6</sup> Among mother-infant pairs followed for 24 months in the parent cohort.

Most WLHIV (25/28, 89%) were diagnosed before pregnancy, all of whom initiated ART before conception. At enrollment, WLHIV had received a median 3.5 years (interquartile range [IQR] 1.5-6) of ART. Most (n=25, 89%) reported receiving the first-line ART regimen of tenofovir (TDF) + lamivudine (3TC) + efavirenz (EFV) as recommended by the Kenya Ministry of Health during study enrollment (2018 - 2020; [30]), while 3 (11%) were receiving an alternative regimen. No WLHIV received a dolutegravir (DTG)-based ART regimen during pregnancy as rollout of DTG in Kenya began in 2018 and was not yet available to study participants at time of enrollment. Only two (7%) WLHIV switched to TDF+3TC+DTG during the first year postpartum. Median CD4 count at enrollment was 562 (IQR 416-680), and 22 (79%) WLHIV reported receiving cotrimoxazole preventive therapy (CPT) during pregnancy. All CHEU received antiretrovirals (ARVs) and CPT for HIV prophylaxis.

### Probability & Timing of Infant CMV Acquisition

CMV status and timing of acquisition were ascertained for 65/72 infants (90%) by CMV DNA PCR and/or serology, of which 47 (72%) acquired CMV and 18 (28%) had no evidence of infection by 12 months of life. Serology results at the end of the study were available for 55/72 infants (76%), and sustained shedding was detected in all but 3 of the antibody positive infants; for each of these 3 infants, serology testing was performed after discontinuation of weekly sampling. There was no significant difference between CHEU and CHU in the mean maximum viral load shed in urine (5.44 vs 4.99 log, p=0.2) or oral swabs (8.29 vs 7.60 log, p=0.07). The 7 infants excluded from further analyses were: one CHU with congenital CMV infection, based on detectable CMV DNA in the first urine sample collected four days after birth followed by sustained shedding, and six infants (1 CHEU, 5 CHU) with <9 months of follow-up or insufficient samples to determine or confirm the timing of infection (Figure S1).

The cumulative probability of CMV acquisition in the first year of life was 72% and did not differ significantly by HIV exposure (CHEU 74% vs CHU 71%, log-rank p=0.653; Figure 1). Median age at primary infection was similar between CHEU (1.7 months, IQR 1.5-2.8) and CHU (2.0 months, IQR 1.4-2.7). Though CHEU appeared to have higher probability of CMV acquisition at several points during follow-up (Figure 1), there was no evidence that the Cox proportional hazards regression model for HIV exposure violated the proportional hazards assumption (Schoenfeld residuals test p=0.439). Additionally, we used z-tests to compare the probability of CMV acquisition at each month between months 1 and 6 as an exploratory analysis; there was no significant difference by HIV exposure at any time point (Holm-Bonferroni corrected p>0.05 for all).

**Figure 1.**
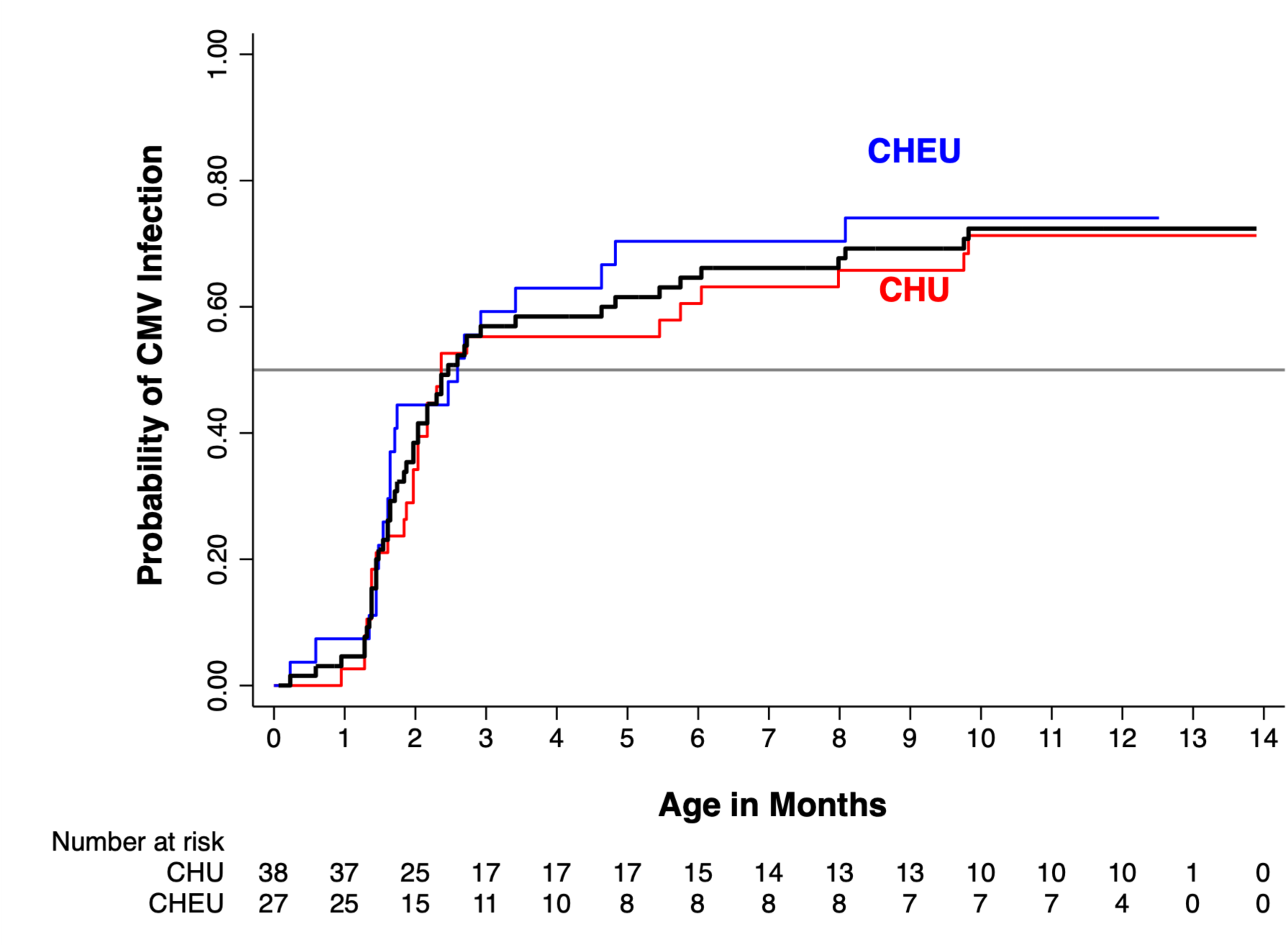
Kaplan-Meier curve for the probability of CMV acquisition. The black line indicates the probability of CMV acquisition among all infants. The horizontal line shows the median survival probability (0.5). Probability did not differ between children who were HIV exposed, uninfected (CHEU, blue) and children who were HIV-unexposed, uninfected (CHU, red; log-rank p=0.653).

### Correlates of CMV Acquisition

Overall incidence of CMV acquisition was 13.5 per 100 person-months (95% confidence interval [CI]: 10.1-18.0) and was similar between CHEU (15.0 per 100 person-months, 95% CI: 9.7, 23.3) and CHU (12.5 per 100 person-months, 95% CI: 8.6, 18.3; Table 2).

**Table 2.**
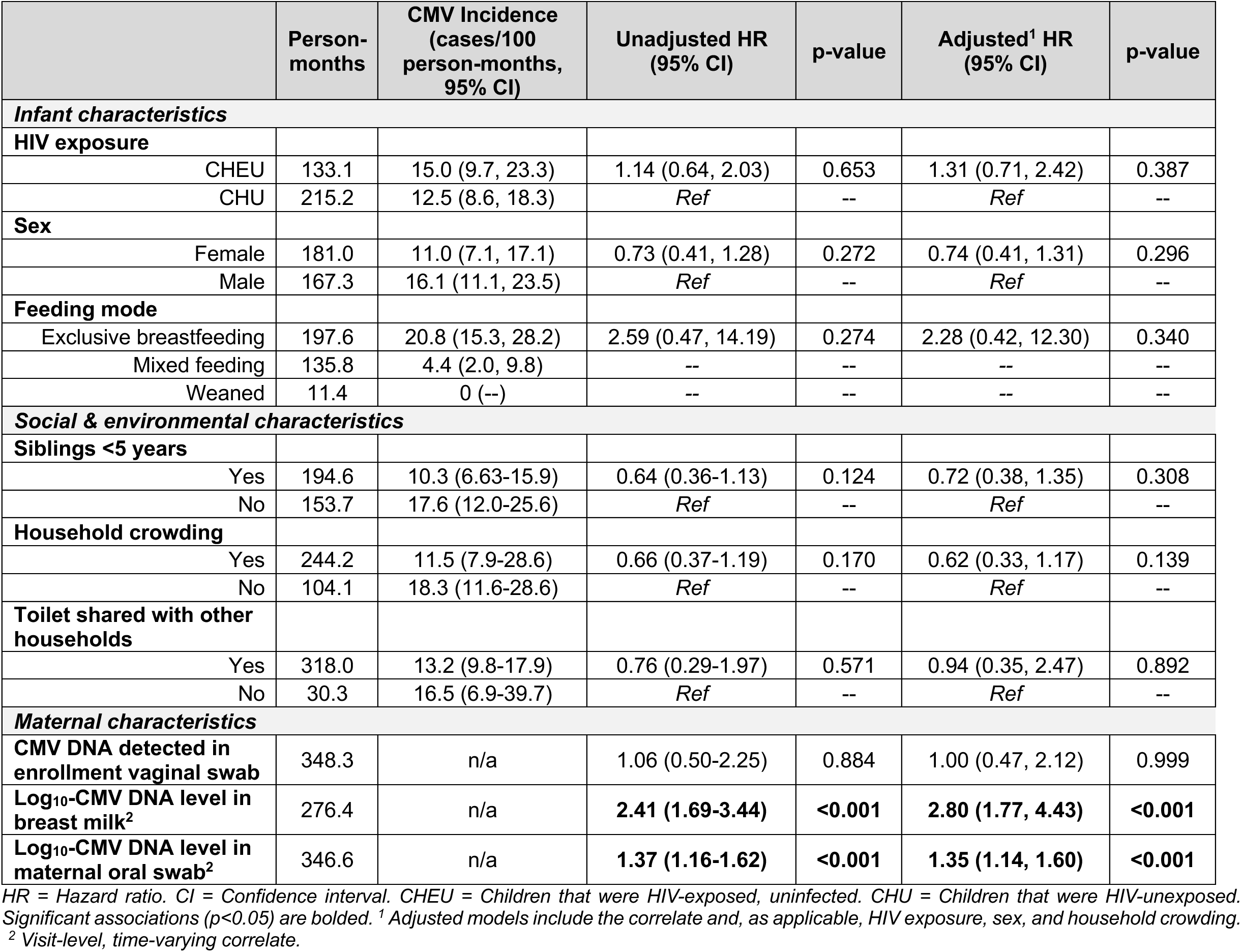
Incidence and hazard ratios for hypothesized correlates of CMV acquisition.

Of the 47 CMV infections, 41 (87%) occurred while infants were EBF, 6 (13%) occurred while infants were receiving mixed feeds, and none occurred after infants were weaned (Table 2). Incidence of CMV acquisition was higher during EBF (20.8 per 100 person-months, 95% CI: 15.3-28.2) than during mixed feeding (4.4 per 100 person-months, 95% CI: 2.0-9.8). The hazard of CMV acquisition did not differ significantly during EBF versus mixed feeding (adjusted hazard ratio [aHR]=2.59, 95% CI: 0.47, 14.19; p=0.274), though because only six infections occurred while infants were not EBF, there was limited statistical power to accurately estimate the hazard of CMV infection associated with feeding mode. Additionally, there was insufficient statistical power to compare the hazard of CMV acquisition associated with no breastfeeding.

Due to high adherence to the recommendations to EBF for 6 months and continued breastfeeding through the first year of life (Table 1), feeding mode was collinear with age in this cohort. We observed similar hazard ratio estimates when adjusting versus not adjusting for feeding mode, and model fit was not significantly improved for any model when including feeding mode as a confounder (likelihood ratio p>0.05 for all; Table S1). Given that age is accounted for in Cox proportional hazards regression and that nearly all CMV infections occurred during EBF, limiting statistical power, we did not additionally adjust for feeding mode in the remaining analyses.

Few characteristics were associated with CMV acquisition (Table 2). We did not observe a significant relationship between HIV exposure and CMV acquisition (aHR=1.31, 95% CI: 0.71, 2.42; p=0.387). All women had detectable CMV DNA in at least one breast milk sample, including women whose infant did not have CMV detection during the period of observation. Each log_10_ increase in breast milk CMV DNA was associated with a nearly three-fold increase in hazard of CMV acquisition in infants (aHR=2.80, 95% CI: 1.77, 4.43; p<0.001). Similarly, there was a 1.35 times greater hazard of CMV acquisition (95% CI: 1.14, 1.60; p<0.001) for every log_10_ increase in maternal oral swab CMV DNA level.

To evaluate whether risk factors for CMV acquisition differed by HIV exposure, we repeated the regression analysis for CHEU and CHU separately (Table S2). Like the full analysis, having siblings <5 years old, household crowding, and sharing a toilet with other households were not significantly associated with CMV acquisition among either CHEU or CHU. There was evidence that HIV exposure modified the association between infant sex and CMV acquisition— among CHU, female infants had 71% lower hazard (aHR=0.29, 95% CI: 0.13, 0.67; p=0.004), while there was no significant difference between male and female CHEU (aHR=2.01, 95% CI: 0.79, 5.10; p=0.142).

Mean log_10_ CMV DNA levels in breast milk were higher among WLHIV than women without HIV (4.17-log vs 3.12-log, p<0.001; Figure 2A), and breast milk CMV DNA level was significantly associated with infant CMV acquisition among both CHEU (aHR=1.70, 95% CI: 1.06, 2.72; p=0.028) and CHU (aHR=4.02, 95% CI: 1.90, 8.55; p<0.001). Mean log_10_ CMV DNA levels were also higher in oral swabs from WLHIV (1.10 vs 0.78 log, p<0.001; Figure 2B). The association between maternal oral swab CMV level and infant CMV acquisition was significant among CHU (aHR=1.50, 95% CI: 1.12, 2.02; p=0.007) and was a trend among CHEU (aHR=1.24, 95% CI: 0.97, 1.57); p=0.085). Most women (60, 83%) did not have detectable CMV DNA in the enrollment vaginal swab, but among women with detectable CMV, mean vaginal log_10_-DNA levels did not differ by HIV status (2.31 vs 2.71 log, p=0.264). As in the full analysis, there was no association between detectable CMV DNA in the enrollment vaginal swab and CMV acquisition for either CHEU (aHR=0.91, 95% CI: 0.35, 2.38; p=0.845) or CHU (aHR=0.93, 95% CI: 0.36, 2.39; p=0.875).

**Figure 2.**
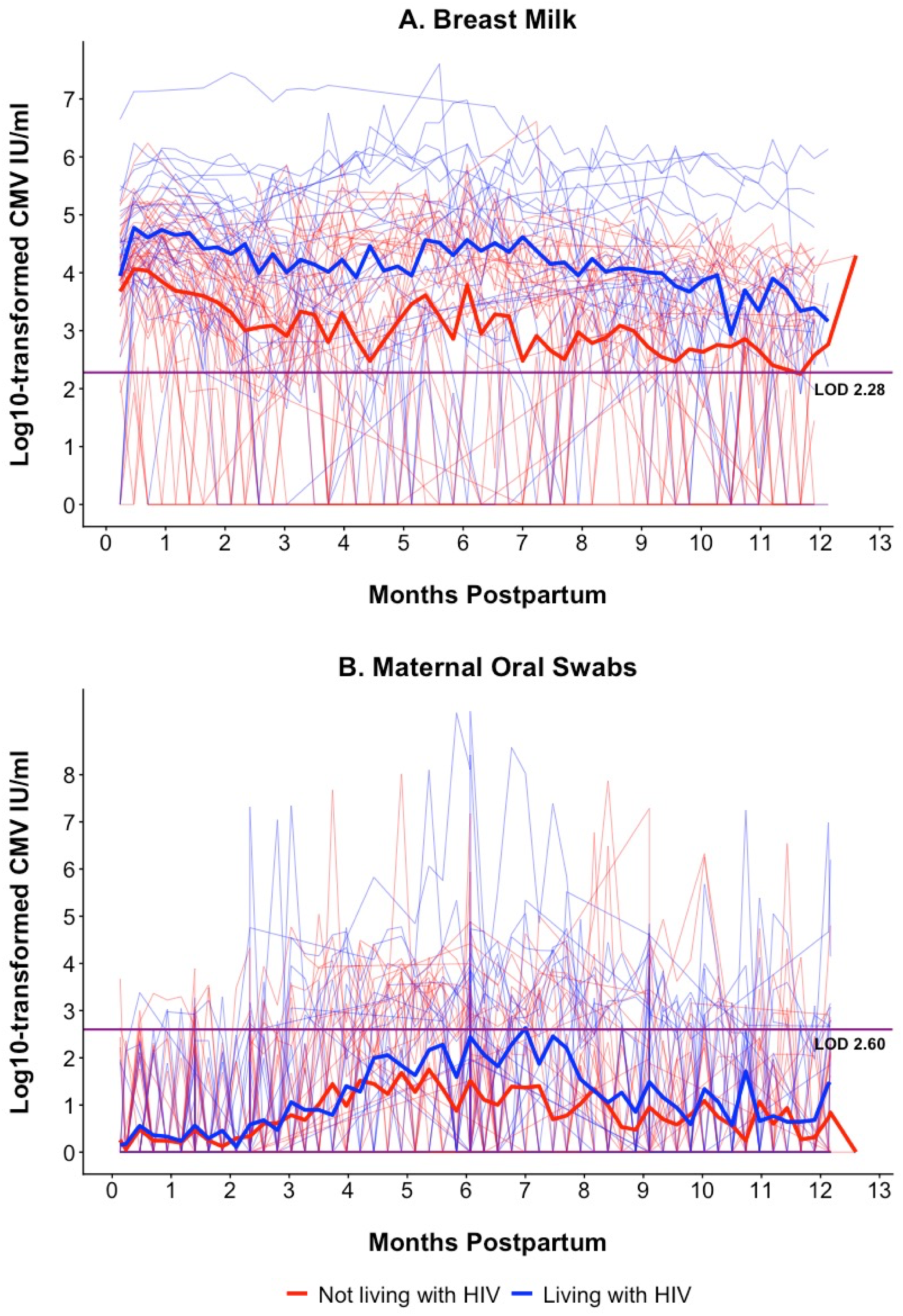
Log_10_-CMV DNA levels in A) breast milk and B) oral swab samples by maternal HIV status. The bold lines represent overall mean log_10_-CMV DNA levels for women living with HIV (blue) and women without HIV (red). The background lines represent individual longitudinal patterns of log_10_-CMV DNA levels. The quantitative PCR limit of detection (LOD) is indicated by the horizontal line in each plot.

Finally, we assessed HIV-associated correlates of CMV acquisition among CHEU (Table 3). There was no association between maternal third trimester (enrollment) CD4 count, ART regimen, or years on ART and infant CMV acquisition. However, infants with postnatal exposure to the recommended first line ART regimens (TDF+3TC+EFV or TDF+3TC+DTG) had a 63% lower hazard of CMV acquisition than infants exposed to alternative regimens (aHR=0.37, 95% CI: 0.15, 0.93; p=0.035). Infant cotrimoxazole prophylaxis also was associated with an 81% lower hazard of CMV acquisition (aHR=0.19, 95% CI: 0.074, 0.47; p<0.001); because CHEU are eligible for cotrimoxazole preventive therapy by 6 weeks of age, this analysis only included infants ages ≥6 weeks who had not yet acquired CMV. All CHEU received ARV prophylaxis as the standard of care for the prevention of mother-to-child HIV transmission, so we could not assess the effect of ARV exposure in this cohort.

**Table 3.** Incidence and hazard ratios for hypothesized HIV-associated correlates of CMV acquisition among CHEU.

|  | Person-months | CMV Incidence (cases/100 person-months, 95% CI) | Unadjusted HR (95% CI) | p-value | Adjusted<sup>1</sup> HR (95% CI) | p-value |
| --- | --- | --- | --- | --- | --- | --- |
| **Maternal CD4 count at enrollment (cells/μl)** | 133.1 | n/a | 1.00 (0.999, 1.00) | 0.619 | 1.00 (0.999, 1.00) | 0.768 |
| **Maternal ART regimen at enrollment<sup>2</sup>** |  |  |  |  |  |  |
| Recommended first line regimen | 127.0 | 13.4 (8.3, 21.5) | 0.52 (0.27, 1.01) | 0.054 | 0.54 (0.25, 1.15) | 0.110 |
| Other regimen | 6.0 | 49.6 (16.0, 153.9) | *Ref* | -- | *Ref* | -- |
| **Maternal years on ART at enrollment** | 133.1 | n/a | 1.05 (0.92, 1.19) | 0.479 | 1.03 (0.90, 1.18) | 0.679 |
| **Maternal postpartum ART regimen<sup>3,4</sup>** |  |  |  |  |  |  |
| Recommended first line regimen | 125.8 | 14.3 (9.0, 22.7) | **0.38 (0.15, 0.96)** | **0.041** | **0.37 (0.15, 0.93)** | **0.035** |
| Other regimen | 5.6 | 35.6 (8.9, 142.3) | *Ref* | -- | *Ref* | -- |
| **Infant cotrimoxazole prophylaxis<sup>4,5</sup>** |  |  |  |  |  |  |
| Yes | 93.9 | 10.6 (5.7, 19.8) | **0.31 (0.10, 0.95)** | **0.040** | **0.19 (0.074, 0.47)** | **<0.001** |
| No | 8.6 | 81.0 (38.6, 169.9) | *Ref* | -- | *Ref* | -- |

HR = Hazard ratio. CI = Confidence interval. Significant associations ( $p < 0.05$ ) are bolded. <sup>1</sup> Adjusted models included the correlate, sex, and household crowding.
<sup>2</sup> Recommended first line regimen was TDF+3TC+EFV. Other regimens included AZT+3TC+EFV and AZT+3TC+ATV/r. <sup>3</sup> Recommended first line regimens were TDF+3TC+EFV or TDF+3TC+DTG. Other regimens included AZT+3TC+EFV, AZT+3TC+ATV/r, TDF+3TC+NVP, TDF+FTC+EFV, and TDF+FTC+DTG. <sup>4</sup> Visit-level, time-varying correlate. <sup>5</sup> Because cotrimoxazole preventive therapy for CHEU is recommended to start at 6 weeks of age, models include only visits conducted when infants were $\geq 6$ weeks of age.

## DISCUSSION

In this cohort of healthy infants, weekly evaluation of CMV shedding across multiple mucosal compartments over the first year of life revealed several new insights into factors modulating vertical CMV transmission. Despite ART initiated before or in early pregnancy, WLHIV had higher breast milk and oral swab CMV DNA levels throughout the postpartum period than women without HIV. While higher CMV levels in these compartments were associated with CMV acquisition among all infants, the hazard of CMV acquisition did not differ significantly between CHEU and CHU, suggesting that additional factors modulate CMV transmission risk. Among CHEU, there was a lower hazard of CMV acquisition when infants received cotrimoxazole prophylaxis and when their mothers received recommended first line ART regimens (TDF+3TC+EFV or DTG) versus alternative first line or second line regimens postpartum. Together, these data suggest that risk of CMV infection among CHEU is modulated by a complex array of virologic, pharmacologic, and immunologic factors.

Few studies have compared postnatal CMV acquisition between CHEU and CHU. We observed that over half of infants acquired CMV by 3 months and the overall probability of CMV acquisition in the first year of life was high (72%). This probability was higher than the 59% observed overall among CHEU and CHU in a similar study of mother-infant pairs in Uganda followed between 2008-2009 with lower rates of breastfeeding [9], but more similar to a study in Zimbabwe during the Option B+ era, in which 74% of CHEU and 48% of CHU acquired CMV by 3 months [31]. In contrast to our findings, a recent study in Cameroon observed higher CMV prevalence at 6 months of age among CHU than CHEU born to WLHIV predominantly receiving TDF+3TC+EFV (61% vs 43%, respectively) [32]. However, like our study, these findings also suggest risk of CMV infection among CHEU has decreased from the pre-ART era.

Vertical CMV transmission declines with improved HIV viral suppression and maternal immune function from ART [33]. In the pre-ART and early ART eras in Kenya, we observed a high incidence of CMV infection among CHEU. In those studies, 90% of CHEU born to women receiving short-course zidovudine during pregnancy acquired CMV by 3 months [34] and 94% of CHEU born to women receiving zidovudine versus 75% of CHEU born to women receiving triple-combination ART acquired CMV by 12 months [33]. Results from our current study with contemporary ART parallel those of the combination ART arm of our previous study, underscoring the importance of maternal ART in modulating vertical CMV transmission among CHEU.

Breast milk is the major route of vertical CMV transmission [35] and breast milk CMV DNA levels were significantly associated with CMV acquisition among all infants, consistent with other studies of CHEU [9,33,36–38] and CHU [11,39–41]. All mothers, regardless of HIV status, had ≥1 breast milk sample with detectable CMV DNA. Among WLHIV, higher HIV viral load and lower CD4 count are associated with greater CMV shedding and earlier and higher risk of CMV infection among CHEU [33,36–38,42]. Despite the fact that most WLHIV initiated ART pre-conception and had CD4 count >200 cells/µl in late pregnancy, we still observed that they had higher breast milk and oral swab CMV DNA levels than women without HIV. Our results are similar to a study in Malawi that found persistently high breast milk CMV DNA levels throughout the first year postpartum among women who started ART during pregnancy and high probability of CMV acquisition among their CHEU despite normal CD4 counts at delivery [43].

Surprisingly, however, risk of CMV acquisition did not differ between CHEU and CHU in our study. Among CHEU, hazard of CMV acquisition was lower when mothers received TDF+3TC+EFV or DTG during pregnancy and postpartum—as recommended by the Kenya Ministry of Health during the study period for first-line ART [30,44]—versus an alternative regimen. Compared to older ART, modern ART such as TDF+3TC+EFV or DTG more effectively reduce HIV viral replication, immune dysfunction, and inflammation. These maternal factors are associated with greater immune dysfunction, inflammation, and morbidity among CHEU, which in turn increase risk of CMV acquisition [45]. It is possible these recommended first-line ART regimens reduced the risk of vertical CMV transmission from WLHIV by improving maternal immune function and/or reduced CHEU’s susceptibility to CMV infection. However, we did not have data to confirm why women received alternative ART regimens or their postpartum HIV viral load or CD4 counts, so it is also possible that this association was confounded by underlying maternal health characteristics or virologic monitoring warranting administration of an alternative regimen. Additional research is needed to determine whether current ART regimens directly influence vertical CMV transmission.

The hazard of CMV acquisition was also lower when CHEU received cotrimoxazole prophylaxis, which is recommended from 6 weeks of age until complete cessation of breastfeeding [30]. To our knowledge, no study has assessed the relationship between cotrimoxazole and CMV incidence among CHEU. As an antibiotic, cotrimoxazole does not directly act against CMV, but it could lower infants’ susceptibility to CMV by preventing or eliminating other infections and related inflammation. It is also possible that infants who did not receive cotrimoxazole as recommended had other underlying sociodemographic or health characteristics that increased their risk of CMV. Further study, ideally in the setting of randomized trials to minimize confounding by other factors, is needed to clarify the effect of cotrimoxazole on CMV acquisition.

Finally, HIV exposure modified the association between infant sex and CMV acquisition— female CHU had a lower hazard of CMV acquisition than male infants, while there was a trend for female CHEU to have a greater hazard of CMV acquisition. Male children have higher levels of inflammation and greater susceptibility to infectious pathogens in childhood, which may explain the lower risk of CMV among female CHU [46]. In a cohort of Kenyan CHEU, female infants were more likely to have HIV-specific T cell responses at age 3 months, which suggests sex-specific immune responses to HIV exposure are possible and could alter CMV risk among female CHEU [47]. However, further investigation is needed to understand mechanisms for this finding.

A limitation of this study is its small sample size. We identified only one case of congenital CMV and therefore could not assess risk factors or compare prevalence by HIV exposure. However, statistical power for our primary analyses was improved by weekly data collection for one year postpartum. This frequency of sampling allowed us to precisely estimate timing of CMV acquisition, which is a unique strength of this study. Additionally, due to parent cohort enrollment criteria, there was low heterogeneity of infant feeding practices, economic indicators, and HIV-associated characteristics. Though we did not adjust for infant feeding in our models, we do not anticipate substantial residual confounding by feeding mode since all CMV infections occurred while infants were still breastfeeding. However, our findings may not be generalizable to populations of infants that are not breastfed. We did not have data for HIV viral load among WLHIV, and so could not include this factor in analyses. Similar third trimester CD4 counts and ART history among WLHIV also may have limited our ability to observe associations between these factors and CMV acquisition among CHEU and these analyses should be repeated in larger studies.

## Conclusion

In the context of universal modern ART, we observed no association between HIV exposure and the probability of CMV acquisition during infancy, and a strong positive association between breast milk CMV DNA level and risk of transmission to the nursing infant. Studies in cohorts with greater heterogeneity of sociodemographic and HIV-associated characteristics are needed to assess whether these findings are consistent at a population level.

## Data Availability

All data produced in the present study are available upon reasonable request to the authors

## ACKNOWLEDGEMENTS

We thank the Linda Kizazi Study participants and their families, our study staff, and the staff and leadership at Mathare North Health Centre for their time and effort that made this study possible. We extend our thanks to the community advisory board members who helped optimize and provide context for our study. We also thank the University of Nairobi and Kenyatta National Hospital for supporting this research.

