## Supplementary Data for "Postnatal CMV acquisition risk is not increased among children who are HIV-exposed, uninfected versus HIV-unexposed in the setting of optimized maternal ART"

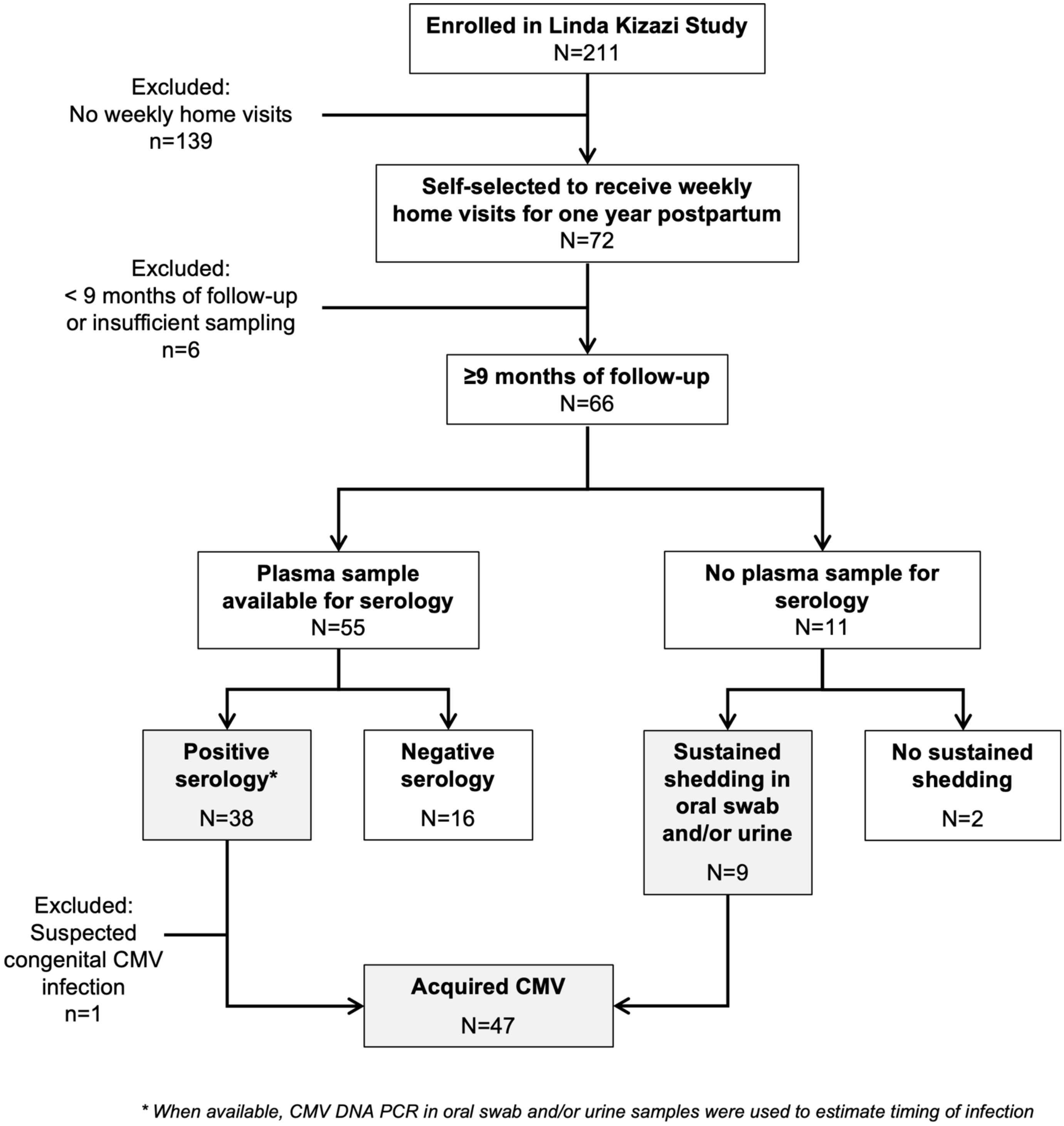


**Figure S1. Flowchart of participant inclusion.** Shaded gray boxes indicate participants with CMV acquisition.

**Table S1. Hazard ratios for hypothesized correlates of CMV acquisition, adjusted for feeding mode.**

| | Adjusted[1] HR (95% CI) | HR p-value | Likelihood Ratio Test[2] p-value |
|---|---|---|---|
| ***Infant characteristics*** | | | |
| **HIV exposure** | | | |
| CHEU | 1.30 (0.70, 2.41) | 0.404 | 0.378 |
| CHU | *Ref* | -- | -- |
| **Sex** | | | |
| Female | 0.74 (0.41, 1.35) | 0.330 | 0.362 |
| Male | *Ref* | -- | -- |
| ***Social & environmental characteristics*** | | | |
| **Siblings <5 years** | | | |
| Yes | 0.71 (0.40, 1.29) | 0.261 | 0.197 |
| No | *Ref* | -- | -- |
| **Household crowding** | | | |
| Yes | 0.68 (0.37, 1.25) | 0.216 | 0.362 |
| No | *Ref* | -- | -- |
| **Toilet shared with other households** | | | |
| Yes | 0.90 (0.36, 2.29) | 0.830 | 0.151 |
| No | *Ref* | -- | -- |
| ***Maternal characteristics*** | | | |
| **$Log_{10}$-CMV DNA level in breast milk** | **2.47 (1.72, 3.55)** | **<0.001** | 0.577 |
| **$Log_{10}$-CMV DNA level in maternal oral swab** | **1.49 (1.24, 1.79)** | **<0.001** | 0.072 |
| **CMV DNA detected in enrollment vaginal swab** | 0.95 (0.44, 2.03) | 0.886 | 0.359 |
| ***HIV-associated correlates[3]*** | | | |
| **Maternal CD4 count at enrollment (cells/µl)** | 1.00 (>0.999, 1.00) | 0.560 | 0.424 |
| **Maternal ART regimen at enrollment** | | | |
| Recommended first line regimen | 0.53 (0.24, 1.15) | 0.107 | 0.411 |
| Other regimen | *Ref* | -- | -- |
| **Maternal years on ART at enrollment** | 1.01 (0.88, 1.16) | 0.843 | 0.197 |
| **Maternal postpartum ART regimen** | | | |
| Recommended first line regimen | **0.37 (0.14, 0.92)** | **0.033** | 0.412 |
| Other regimen | *Ref* | -- | -- |
| **Infant cotrimoxazole prophylaxis** | | | |
| Yes | **0.21 (0.078, 0.56)** | **0.002** | n/a[4] |
| No | *Ref* | -- | -- |

*HR = Hazard ratio. CI = Confidence interval. CHEU = Children that were HIV-exposed, uninfected. CHU = Children that were HIV-unexposed. Significant associations (p<0.05) are bolded. [1] Adjusted models include the correlate, feeding mode (exclusive breastfeeding, mixed feeding, weaned), and, as applicable, HIV exposure, sex, and household crowding. [2] Likelihood ratio test comparing adjusted models with feeding mode to adjusted models without feeding mode. [3] Among CHEU. [4] Insufficient power; there were no infants that did not receive cotrimoxazole during mixed feeding.*

**Table S2. Hazard ratios (HR) for hypothesized correlates of CMV acquisition among CHEU and CHUU.**

| | HIV-unexposed, uninfected | | | | HIV-exposed, uninfected | | | |
|---|---|---|---|---|---|---|---|---|
| | **Unadjusted HR (95% CI)** | **p-value** | **Adjusted[1] HR (95% CI)** | **p-value** | **Unadjusted HR (95% CI)** | **p-value** | **Adjusted[1] HR (95% CI)** | **p-value** |
| ***Infant characteristics*** | | | | | | | | |
| **Feeding mode** | | | | | | | | |
| Exclusive breastfeeding | 2.19 (0.20, 23.72) | 0.518 | 1.56 (0.15, 16.06) | 0.708 | n/a[2] | -- | n/a[2] | -- |
| Mixed feeding | *Ref* | -- | *Ref* | -- | *Ref* | -- | *Ref* | -- |
| **Sex assigned at birth** | | | | | | | | |
| Female | **0.29 (0.12, 0.66)** | **0.003** | **0.29 (0.13, 0.67)** | **0.004** | 2.08 (0.80, 5.40) | 0.135 | 2.01 (0.79, 5.10) | 0.142 |
| Male | *Ref* | -- | *Ref* | -- | *Ref* | -- | *Ref* | -- |
| ***Social & environmental characteristics*** | | | | | | | | |
| **Siblings <5 years** | | | | | | | | |
| Yes | 0.74 (0.34, 1.58) | 0.429 | 0.71 (0.34, 1.50) | 0.375 | 0.53 (0.21, 1.34) | 0.182 | 0.52 (0.19, 1.43) | 0.206 |
| No | *Ref* | -- | *Ref* | -- | *Ref* | -- | *Ref* | -- |
| **Household crowding** | | | | | | | | |
| Yes | 0.72 (0.34, 1.52) | 0.392 | 0.81 (0.37, 1.76) | 0.590 | 0.52 (0.19, 1.48) | 0.223 | 0.55 (0.18, 1.69) | 0.294 |
| No | *Ref* | -- | *Ref* | -- | *Ref* | -- | *Ref* | -- |
| **Toilet shared with other households** | | | | | | | | |
| Yes | 0.58 (0.26, 1.29) | 0.181 | 0.95 (0.39, 2.29) | 0.902 | 1.36 (0.084, 21.98) | 0.829 | 5.56 (0.26, 119.15) | 0.272 |
| No | *Ref* | -- | *Ref* | -- | *Ref* | -- | *Ref* | -- |
| ***Maternal characteristics*** | | | | | | | | |
| **$Log_{10}$-transformed CMV viral load in breast milk** | **4.20 (1.92, 9.20)** | **<0.001** | **4.02 (1.90, 8.55)** | **<0.001** | **1.90 (1.26, 2.87)** | **0.002** | **1.70 (1.06, 2.72)** | **0.028** |
| **$Log_{10}$-transformed CMV viral load in oral swabs** | **1.48 (1.12, 1.97)** | **0.006** | **1.50 (1.12, 2.02)** | **0.007** | 1.23 (0.99, 1.54) | 0.058 | 1.24 (0.97, 1.57) | 0.085 |
| **CMV DNA detected in enrollment vaginal swab** | 1.04 (0.40, 2.69) | 0.936 | 0.91 (0.35, 2.38) | 0.845 | 0.94 (0.31, 2.84) | 0.911 | 0.93 (0.36, 2.39) | 0.875 |

*HR = Hazard ratio. CI = Confidence interval. Significant associations ($p<0.05$) are bolded. [1] Adjusted models include the correlate and, as applicable, sex and household crowding.*
*[2] Insufficient power to estimate hazard ratio.*
